# Robotic versus Electromagnetic Bronchoscopy for Pulmonary LesIon AssessmeNT using integrated intraprocedural imaging: study protocol for the RELIANT 2 trial

**DOI:** 10.1101/2025.06.19.25329969

**Authors:** Rafael Paez, Ankush P. Ratwani, Jennifer D. Duke, Greta Bridwell, Kaele Leonard, James M Katsis, Edward M Pickering, Jeffrey Thiboutot, Heidi Chen, Sheau-Chiann Chen, Samira Shojaee, Lonny Yarmus, Robert J. Lentz, Fabien Maldonado, the Interventional Pulmonary Outcomes Group

## Abstract

**Background:** Robotic assisted bronchoscopy (RAB) and electromagnetic navigational bronchoscopy (ENB) are two approaches to biopsy peripheral pulmonary lesions (PPLs). A recently completed cluster randomized controlled trial, RELIANT (NCT05705544), showed no difference in diagnostic yield between these two modalities. The ENB platform used in RELIANT included integrated digital tomosynthesis, allowing for real-time assessment of PPL location for more precise targeting at the time of biopsy. RAB has since been integrated with cone beam computed tomography (CBCT) to accomplish the same intraprocedural correction. It is unclear if the diagnostic yield of RAB with integrated CBCT (RAB-CBCT) is superior to ENB with integrated digital tomosynthesis (ENB-DT).

**Methods:** **R**obotic versus **E**lectromagnetic Bronchoscopy for Pulmonary **L**es**I**on **A**ssessme**NT** using integrated intraprocedural imaging (RELIANT 2) is an investigator-initiated, multicenter, open label, superiority, cluster randomized trial. At each participating institution, procedural rooms are randomly assigned to either RAB-CBCT or ENB-DT, with each procedure room-day considered a cluster. All adult patients undergoing navigational bronchoscopy for evaluation of PPL(s) are eligible. Allocation is concealed from schedulers, proceduralists, and patients until the morning of procedure when each room is randomized to a platform. The primary endpoint is the diagnostic yield, defined as the proportion of cases yielding a specific benign or malignant diagnosis per current ATS/ACCP definition. Secondary and safety endpoints include procedure duration and procedural complications. Enrollment began on November 11, 2024 and is expected to enroll approximately 440 patients in 220 clusters.

**Discussion:** RELIANT 2 is an ongoing cluster randomized trial comparing the diagnostic yield of RAB-CBCT to that of ENB-DT in patients undergoing bronchoscopy to biopsy PPLs. This trial will help address some of the limitations of the recently published RELIANT trial.

**Trial Registration:** The trial was registered in ClinicalTrials.gov (NCT06654271) on October 21, 2024, prior to patient enrollment.

## Background

Each year, millions of pulmonary nodules are identified during routine imaging (1). While most are benign, some represent malignancy, necessitating a timely and accurate diagnosis. Given the small size and peripheral anatomical location of many of these lesions, advanced navigational systems, such as electromagnetic navigation bronchoscopy (ENB) and robotic-assisted bronchoscopy (RAB), are often used (2,3).

ENB-DT uses an electromagnetic field to track and guide a navigation catheter toward the lesion of interest. The Illumisite platform (Medtronic, Minneapolis, MN, U.S.) includes integrated digital tomosynthesis, which provides real time intraprocedural imaging and allows the operator to correct for computed tomography (CT)-to-body divergence by updating the location of the lesion. ENB-DT has demonstrated improved diagnostic performance compared to ENB without integrated DT (4,5). The Ion Endoluminal system (Intuitive Surgical, Sunnyvale, CA, U.S.), one of three available RAB platforms, uses shape-sensing technology to track the location of the catheter in the airways with improved precision and stability during navigation and biopsy. Although originally used with two-dimensional fluoroscopy, the use of cone-beam computed tomography (CBCT) has been widely adopted as it allows the proceduralist to adjust the position of the catheter relative to that of the nodule guided by intraprocedural three-dimensional (3D) imaging (6).

Despite their widespread use, there is limited data comparing these two commonly used bronchoscopic modalities. The RELIANT randomized trial partially addressed this gap by comparing RAB without integrated CBCT to ENB-DT and showed similar diagnostic yield and no significant difference in the rate of complications (7,8). However, RELIANT was conducted at a single academic medical center with experienced operators, which may limit the generalizability of the results. Additionally, while CBCT was used with RAB in RELIANT, intraprocedural images were not transmitted to the navigational platform to update the target location, and catheter adjustments were instead manually estimated by the bronchoscopist. Recently, CBCT was integrated with RAB (RAB-CBCT) allowing the proceduralist to update the position of the nodule in the targeting system for precise, mid-procedure adjustments rather than by manual estimation. This recent integration is believed to increase the diagnostic yield of RAB-CBCT (9).

To address these limitations, we have designed and are conducting RELIANT 2, a multicenter cluster randomized controlled trial that will test the hypothesis that the diagnostic yield of RAB-CBCT is superior to that of ENB-DT in patients undergoing advanced diagnostic bronchoscopy for PPL biopsy.

## Methods

### Trial Design

RELIANT 2 is an investigator-initiated, multicenter, open label, superiority, cluster randomized, controlled trial comparing the diagnostic yield of RAB-CBCT to that of ENB-DT in patients undergoing bronchoscopy to biopsy a PPL. The trial is being conducted at three academic medical centers in the United States. Consistent with the pragmatic clinical trials concept (10,11), this study is embedded within routine clinical workflow and has broad eligibility criteria. The only aspect of clinical care influenced by the protocol is the device used each day. Similar to RELIANT (7,8), a cluster level design was chosen due to logistic constraints: while both RAB-CBCT and ENB-DT are mobile, they require significant time and effort to set up, and all three sites perform multiple bronchoscopic biopsies each day with one site concurrently using two operating rooms, each using one of the two navigational platforms. Thus, each platform is set up in one procedure room and used for every patient undergoing biopsy in that room on that day as per standard of care. This study has been approved by the Institutional Review Boards at Vanderbilt University Medical Center (IRB 241103), Rush University Medical Center (IRB 24110802) and Johns Hopkins University (IRB 00473086). The trial was registered on ClinicalTrials.gov (NCT06654271) prior to the study opening.

### Study Setting

The trial is being conducted at Vanderbilt University Medical Center (VUMC), Johns Hopkins University (JHU) and Rush University Medical Centers (Rush), which are tertiary academic medical centers with expertise in advanced bronchoscopic procedures. At all three centers, advanced diagnostic bronchoscopy is performed in rooms dedicated to bronchoscopy with full anesthesia support.

### Study Population

All adults 18 years or older scheduled for navigational bronchoscopy for biopsy of a PPL at a participating site are eligible. Patients enrolled in another study requiring the use of a specific bronchoscopic platform and patients who decline to participate are excluded.

### Informed Consent

Given the cluster level design, the device allocation occurs at the start of the day, prior to patients’ arrival in the hospital. A full research informed consent for the use of participants’ data is being obtained from all patients prior to enrollment. The informed consent is obtained by proceduralists or a member of the study team at the time of clinical informed consent for the procedure (7,8).

### Randomization and Blinding

Similar to RELIANT (7,8), parallel cluster randomization is used given the impracticability of patient-level randomization. Each morning, procedure rooms are randomly assigned to use either RAB-CBCT or ENB-DT (1 procedure room-day = 1 cluster). The randomization sequence was generated by a biostatistician not involved in patient care. Randomly permuted blocks of variable size, stratified by site and number of procedure rooms available are used. Allocation is concealed in sealed, opaque envelopes marked with the cluster number, prepared by research staff not involved in patient recruitment or patient scheduling. An envelope allocating the platform assigned for the day is opened the morning of the procedure by bronchoscopy staff preparing the room.

Due to the nature of the interventions, blinding of bronchoscopists and patients through each procedure is not feasible. However, patients are scheduled for their procedure days in advance by a scheduler without knowledge of what platform the procedure room will be randomized to; thus, the patient, the proceduralist, and the scheduler are blinded at the time of scheduling and remain blinded until the day of the procedure. Outcome assessors, including pathologists assessing biopsy specimens are blinded throughout the study.

### Trial Interventions

Procedure room-days assigned to ENB-DT will use the Illumisite™ platform (Medtronic, Minneapolis, MN, U.S.) and those assigned to RAB-CBCT will use the Ion Endoluminal System (Intuitive Surgical, Sunnyvale, CA, U.S.) with CBCT (OEC 3D, GE HealthCare, U.S.) for all navigation bronchoscopies scheduled in that room that day. The only aspect of the procedure influenced by the study is the bronchoscopy platform assignment. ENB-DT and RAB-CBCT procedures are conducted per routine practice as previously described (7, 12, 13). Briefly, all bronchoscopies are performed under general anesthesia with neuromuscular blockade. Radial endobronchial ultrasound is available for all procedures. Biopsy tools including transbronchial needles, biopsy forceps, cryoprobes, and/or other sampling devices are used at the discretion of the bronchoscopist. Rapid on-site evaluation (ROSE) is available to assess specimen adequacy at the discretion of the proceduralist. All patients recover based on routine protocols, which includes two hours of monitoring in the recovery area. Imaging to assess possible pneumothorax is obtained if clinically indicated at the discretion of the proceduralist. Post procedure, patients will be managed and followed per usual care.

### Outcomes

#### Primary Outcome

As in RELIANT (7,8), the primary endpoint is the diagnostic yield, defined as the proportion of procedures that result in a specific benign or malignant diagnosis based on the presence of pathological findings that readily explain the presence of a PPL according to recent consensus statement (14). The following pathological findings are pre-specified as diagnostic:

- Malignancy
- Specific benign pathologic finding including:

- Organizing pneumonia
- Frank purulence/robust neutrophilic inflammation with positive culture data
- Granulomatous inflammation
- Other specific benign findings such as hamartoma, amyloidoma

Biopsies not meeting any of the above lesional histopathological criteria will be adjudicated as non-diagnostic, including biopsies with normal lung parenchyma, atypia not diagnostic of malignancy, or non-specific inflammation. Procedure will also be adjudicated as not meeting the primary outcome if the procedure starts but no biopsies are obtained such as unable to reach the nodule, complication occurs before biopsies are obtained, or equipment failure. Biopsies obtained without the use of guided bronchoscopy (e.g., sampling of central lymph nodes using the linear endobronchial ultrasound bronchoscope) will not be included in the diagnostic yield calculations. Only the index bronchoscopy will be included in the diagnostic yield calculation for patients undergoing multiple bronchoscopies during the study period.

#### Secondary, Safety and Exploratory Outcomes

The secondary outcome is the duration of navigational bronchoscopy defined as time from beginning of registration steps to the removal of the navigation catheter or robotic bronchoscope after completion of the navigation procedure.

The safety outcome is procedural complications within 7 days of the bronchoscopy as defined by the Common Terminology Criteria for Adverse Events (CTCAE v4.0) including pneumothorax; bronchopulmonary haemorrhage requiring additional intervention beyond suctioning and wedging; respiratory failure requiring hospitalization, supplemental oxygen or respiratory support; and anaesthetic complications.

Exploratory clinical outcomes include:

- Radiation exposure, defined as radiation dose delivered to the patient during the study bronchoscopy, recorded as a dose area product (mGy/cm2)
- Additional diagnostic procedures, defined as any diagnostic procedure performed after the study bronchoscopy which targets the same peripheral lesion (including repeat bronchoscopy, transthoracic needle biopsy, or surgical lung biopsy) between completion of the index procedure and 12 months. Repeat biopsies of lesions determined to be malignant by study bronchoscopy which are 1) performed specifically to obtain additional tissue for further testing but that does not change the malignant diagnosis, or 2) have therapeutic intent such as surgical resection, will not be considered additional diagnostic procedures.
- Diagnostic accuracy at 12 months post-biopsy, defined as the percentage of patients with biopsies that showed a specific diagnosis (malignancy or a specific benign condition) that was confirmed to be accurate through 12 months of clinical follow-up (13).
- Best radial endobronchial ultrasound (rEBUS) signature at any point during procedure including concentric, eccentric and no signature.

### Recruitment

All patients undergoing advanced diagnostic bronchoscopy for biopsy of a PPL are screened for eligibility. Patients who do not meet inclusion criteria are considered ‘ineligible.’ Patients who meet inclusion criteria but also meet at least one exclusion criterion are considered ‘excluded.’ For patients who meet inclusion criteria but are not enrolled, the reason for exclusion is recorded. After confirming the patient meets eligibility criteria, the patient is approached for enrollment in the study (**Fig 1**). Written research informed consent is obtained at the time of procedural consent. Participating cites have ample volume to achieve adequate enrollment for this trial as seen in RELIANT (8). Patients are followed per usual care at the discretion of the proceduralist. Standard operating procedures are in place at participating sites to ensure patients return for their routine follow-up.

**Fig 1.**
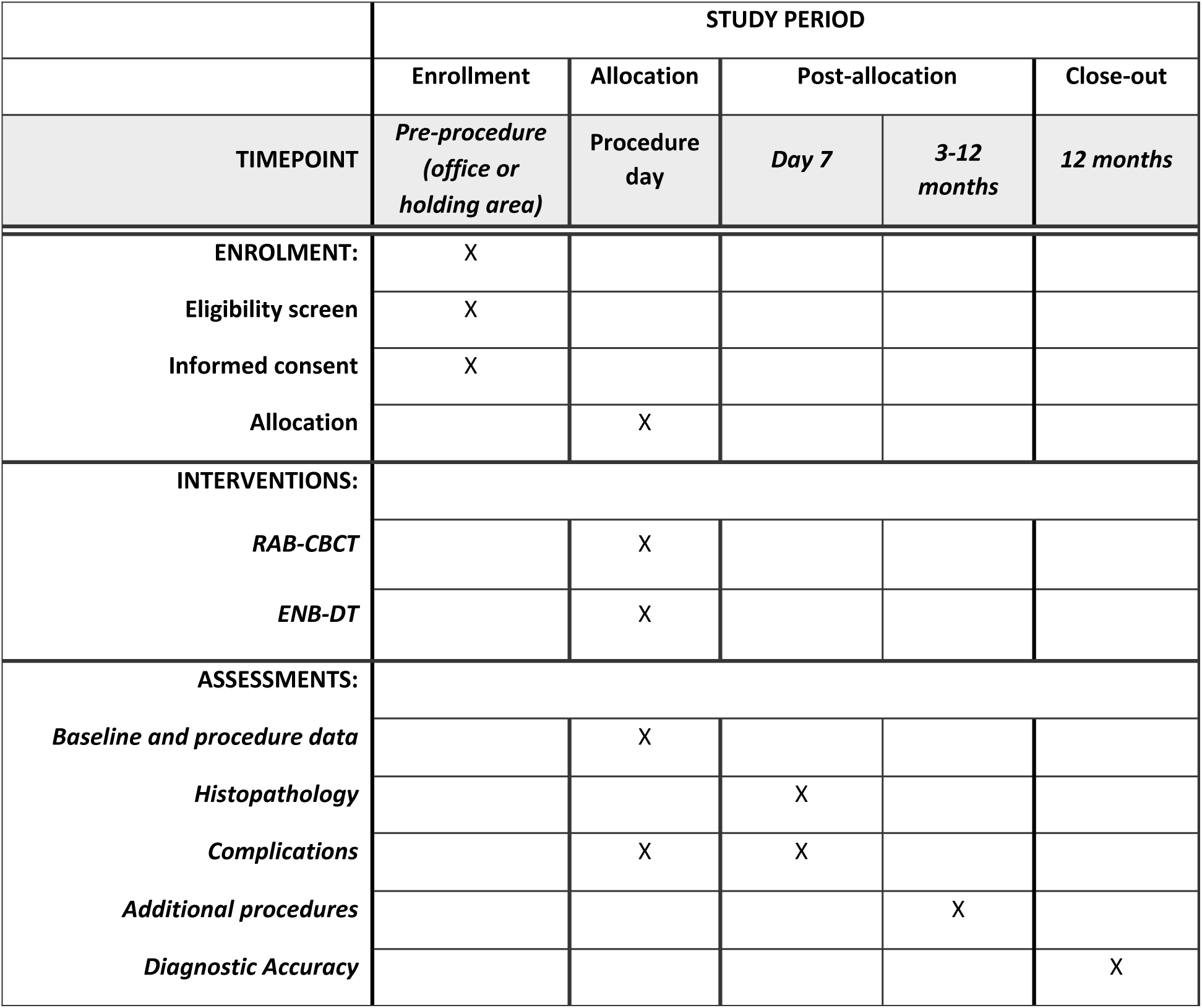
Study Calendar

### Data Collection and Management

In this pragmatic trial, procedural and clinical data recorded in the electronic health record as part of routine clinical care are recorded in a Health Insurance Portability and Accountability Act (HIPAA) compliant REDCap database (15). Data collected includes demographics, medical comorbidities, lesion size and other radiological characteristics, procedure details including duration, platform and tools used, specific histopathologic findings, radiation dose, and any complications that occur during or after the procedure. Follow up data including repeat bronchoscopic biopsy and additional biopsies obtained by alternative means are collected if clinically indicated. Data are entered by investigator-proceduralists and research coordinators experienced with the database, which is password protected and requires two factor authentications if accessed from outside the firewall. Research informed consents will be kept in a research binder in a locked file cabinet in a locked office. Only key study personnel approved by the IRB will have access to this database.

### Data and Safety Monitoring

A data and safety monitoring board (DSMB) was appointed to oversee the trial and review one interim analysis at the midpoint of the study. The DSMB consists of 3 members with expertise appropriate to the conduct of the study, including an interventional pulmonologist, a clinical trialist, and a biostatistician. The DSMB evaluates any research-related serious adverse events (SAEs) or unanticipated adverse events (AEs). The appointment of all members was contingent upon the absence of any conflicts of interest. The DSMB developed a charter and reviewed the protocol and appropriate regulatory documents during its first meeting. The DSMB will meet at approximately 50% enrollment to review interim analysis and safety events. The DSMB can recommend that the trial ends, be modified, or continue unchanged based on interim analysis results or safety concerns.

This pragmatic trial compares two bronchoscopic approaches routinely done to biopsy PPLs. Clinical adverse events of bronchoscopy are well described in the literature and discussed in detail with the patient at the time of procedure consent. Thus, these are considered to be related to the clinical procedure rather than related to the research. The main risk of research is breach of privacy and confidentiality, and we have many safeguards in place. Clinical adverse events are systematically collected and reviewed by the study team and DSMB. Serious unanticipated adverse events are reported to the DSMB and IRB per current institutional standards. The RELIANT 2 trial is overseen by a steering committee, which includes principal investigators, and study members. The study team meets weekly to monitor enrollment, adverse events, data quality and completeness, and to discuss any study updates.

### Sample Size Calculation

The diagnostic yield of ENB varies widely in the literature (2,16). Based on data from prior studies and our own published data, we estimated the diagnostic yield of ENB-DT to be 80% (4,12,13). There are limited data on the diagnostic yield of RAB-CBCT (9); however, a 10% increment in diagnostic yield would be considered clinically significant and sufficient to influence practice or hospital capital investment. Assuming the diagnostic yield of ENB is 80% and cluster size of 2, 110 clusters (procedure room-days) per group will provide at least 80% power to detect a 10% increment in diagnostic yield (i.e. 90% diagnostic yield in RAB with integrated CBCT) at a two-sided type I error rate of 5%, with intracluster correlation of 0.1 (based on prior data from RELIANT). Based on an average of 2 subjects per cluster, we will enroll approximately 440 participants in the trial.

### Interim Analysis

This trial will be monitored for efficacy at 50% enrollment (at least 220 evaluable participants, 110 in each arm). The Z statistics of the efficacy boundary at the interim analysis is 2.96 using the O’Brien-Fleming spending function with a corresponding significance level for efficacy of p<0.0015. Because both devices are standard of care interventions to which patients would be exposed regardless of participation in the study and given that even small differences between groups might be clinically meaningful, a futility stopping boundary was not included.

### Statistical Analysis Plan

The statistical analysis plan will follow a similar approach to RELIANT (7,8) and will be conducted using R (version 4.4.0). Baseline demographic and clinical data will be described overall and by group. Categorical variables will be described using frequencies and proportions, and continuous variables will be described using means and standard deviations, as well as medians and interquartile ranges when appropriate. We will describe the outcome variables overall and grouped by study arm using the same approach as for the demographic data. Summary statistics and graphical representations may be displayed, and missingness will be reported for each variable. No statistical comparisons between groups will be made for this descriptive analysis.

#### Primary outcome

All randomized patients who undergo navigational bronchoscopy will be included in the main analysis. Participants will be evaluated by allocated bronchoscopy platform regardless of what platform was used. The primary outcome, diagnostic yield, will be compared between groups using a generalized linear mixed model with two-sided test. The primary model will be covariate-adjusted, including random effects for operator and fixed effects for device assignment, lesion size, density, peripheral location, bronchus sign. Should the model demonstrate signs of overfitting, covariates may be selected based on priority order (device assignment, lesion size, density, peripheral location, bronchus sign, operator).

#### Secondary and exploratory outcome

A sensitivity analysis using a *per protocol* approach will be conducted to analyze participants based on the device used. We will use the same approach as the primary analysis, an adjusted generalized linear mixed effects model using the same covariates.

The secondary outcome, procedure duration, will be compared between study groups using a linear mixed model. If the procedure duration is skewed, alternative models may be pursued. Analysis of the exploratory endpoints will follow a similar approach.

#### Safety analysis

Procedure complications are expected in usual care, although uncommon (8). We will report all procedure complications for each device, overall and by type. If event rates exceed 5%, we may proceed with a comparative analysis, which will involve a generalized linear regression model for binary outcomes as specified for the main analysis, with the exception that covariate adjustment may not be possible. The safety analysis dataset will group participants by device used, regardless of assignment.

All model results will be summarized with point estimates and 95% confidence intervals, which will be emphasized over p-values when reporting the results for secondary outcomes. No adjustments for multiplicity will be made.

#### Differential effects

To determine whether differences in outcomes are dependent on baseline characteristics, we will introduce interaction terms into the models developed for the main analysis. Specifically, we will test the interaction between device assignment and the subgrouping variable. Each variable will be tested one by one, such that all main effects but only one interaction term is included at a time by the likelihood-ratio test. The following putative subgrouping variables are prespecified:

- Nodule size (continuous and grouped as <1.5cm, 1.5-3cm, >3cm)
- Presence of bronchus sign
- Solid vs subsolid nodule
- Peripheral vs central location

Missingness of the primary or secondary outcomes is not expected due to the proximity of its measurement with the procedure and its integration into clinical documentation. Procedures missing the primary outcomes will be considered non-diagnostic. Missing covariates will be imputed using multiple imputation with predictive mean matching. There may be missingness on exploratory outcomes. For missing exploratory outcomes, a complete case analysis will be performed.

### Dissemination plans

After study completion and data analysis, the results will be published in a peer-reviewed journal and will be made available in ClinicalTrials.gov. Authorship will be consistent with the International Committee of Medical Journal Editors (ICMJE) guidelines. The publication of sub-studies and post-hoc analyses will not precede the primary publication. All authors are expected to disclose financial relationships or affiliations that could be considered conflicts of interest per journal or medical society requirements. The protocol and the statistical analysis plan will be available with the published study manuscript. Deidentified participant-level dataset will be available upon reasonable request and with appropriate IRB approval.

## Discussion

RELIANT 2 is an ongoing multicenter, open label, cluster randomized controlled trial comparing the diagnostic yield of RAB-CBCT to that of ENB-DT. We recently completed RELIANT, which showed no significant difference in diagnostic yield between RAB without integrated CBCT and ENB-DT (8). However, some important questions remained. First, the study was conducted at a single academic institution with experienced operators, limiting generalizability. Second, while CBCT was available during RAB in RELIANT, it was only used in 54% of cases and not integrated with the RAB. Accumulating data suggests integrated CBCT improves the diagnostic yield of RAB (9). However, high-quality evidence is needed to confirm these findings and inform patient care and hospital system capital investments (17).

The trial was designed using the same pragmatic framework and infrastructure used in RELIANT (7,8). The study procedures are embedded within routine clinical workflows, which minimizes disruption in clinical care and enhances accrual. Eligibility criteria are broad, co-interventions are at the discretion of proceduralists, and the primary outcome of diagnostic yield is patient-centered (17). Cluster randomization was also used for this trial given impracticability of individual level randomization. This approach reflects real-world practice where the choice of bronchoscopy platform is often driven by logistical constraints rather than clinical criteria. Such a design also mitigates potential biases introduced by individual operator preference. As done for RELIANT, and recommended by society guidelines (13), diagnostic yield is the primary outcome. This patient-centered primary endpoint features immediate adjudication, allowing rapid generation of clinically relevant data that is used to inform patient care while minimizing the burden of long term follow up. Secondary, safety, and exploratory outcomes are also relevant patient centered outcomes and allow assessment of the trade-offs between the two competing technologies. This is particularly relevant in an era of value-based care, where procedural efficiency and diagnostic accuracy must be balanced with patient safety and resource utilization.

If the study demonstrates that RAB-CBCT offers superior diagnostic performance compared to ENB-DT, it could have important implications for institutional technology adoption, training priorities, and procedural planning. Conversely, comparable performance by ENB-DT despite a lower capital investment required and less intensive logistical burden would support its continued use, particularly in resource-constrained settings. The results may also contribute to the evolving guidelines for lung nodule management and may influence future studies investigating bronchoscopic approaches to biopsy pulmonary lesions. Ultimately, the RELIANT 2 trial seeks to deliver actionable evidence to optimize the diagnostic pathway for patients with suspected lung cancer and support the ongoing evolution of interventional pulmonology research.

## Data Availability

Deidentified research data will be made available when the study is completed and published

## Disclosures

Fabien Maldonado: consulting for Medtronic, Intuitive, and J&J. Research funding from Medtronic. Jeffrey Thiboutot: consulting for Verathon, Research funding from Erbe. Lonny Yarmus: consulting from Olympus and Intuitive. Research funding from Erbe and Ambu. Samira Shojaee: Research funding from Cook Medical. Robert Lentz: consulting for Intuitive. Jennifer Duke: consulting for Intuitive, research funding from Cook Medical.

## Funding

This study was in part supported by ATS Research Program Grant number: 23-24D7 to Rafael Paez. The sponsor had no role in the design, conduct, analysis, and reporting of trial.

## Author Contributions

RP, RJL, SS and FM designed the study. RP, RJL, JD, AR, GD, SS, KL, JK, EP, JT, LY, and FM are enrolling and consenting patients. HC, SCC, RP and FM developed biostatistical analysis. All authors contributed to final protocol. RP, and FM wrote the manuscript. All authors reviewed, edited, and approved the final manuscript.

## Notes

### Clinical Trial

ClinicalTrials.gov (NCT06654271)

### Funding Statement

Yes

### Author Declarations

This study has been approved by the Institutional Review Boards at Vanderbilt University Medical Center (IRB 241103), Rush University Medical Center (IRB 24110802) and Johns Hopkins University (IRB 00473086).

